# Diagnostic accuracy of a single-reaction, pan-lineage Lassa virus RT-PCR at two Nigerian laboratories: a retrospective and prospective cohort study

**DOI:** 10.64898/2026.11.03.26367181

**Authors:** Nelson A. Adedosu, Suleiman Y. Alhaji, Johnson Etafo, Idris I. Abduljalail, Ruth I. Aderibigbe, Anita O. Atu, Olufunke Gbenga-Ayeni, Bawa A. Mikail, Ronke I. Uzuajemeh, Aisha M. Yarima, Nancy Njoku-Attah, Iain J. MacLeod

## Abstract

**Background:** Lassa virus (LASV) intra- and inter-lineage nucleotide diversity has made it challenging to develop a single-reaction RT-PCR assay that can detect all viral variants with equal sensitivity. We evaluated PANDAA^®^ LASV, a single-reaction pan-lineage RT-PCR assay, at two Nigerian Lassa fever laboratories against the RealStar Lassa Virus RT-PCR Kit 2.0, the current diagnostic standard in Nigeria, which requires two separate reactions per patient sample to mitigate against false negative results.

**Methodology and Principal Findings:** We conducted a diagnostic accuracy study using retrospectively (*n* = 200) and prospectively (*n* = 100) plasma samples that had been collected from patients meeting the standard case definition of suspected Lassa fever for testing by real-time PCR at Abubakar Tafawa Balewa University Teaching Hospital in Bauchi and Federal Medical Centre in Owo. PANDAA^®^ LASV sensitivity was 98.6% (138/140; 95% CI 94.9–99.8%), specificity 99.4% (155/156; 96.5–100%), and accuracy 99.0% (293/296; 97.1–99.8%). The positive likelihood ratio was 153.8 (21.8–1084.9) and the negative likelihood ratio 0.01 (0.00–0.06). PANDAA Ct values correlated significantly with the reference assay L gene target (Spearman’s *ρ* = 0.80; *p* < 0.0001) and GPC target (*ρ* = 0.69; *p* < 0.0001). Three confirmed discordant results occurred at high Ct values near assay detection limits. Performance was consistent across sites with overlapping confidence intervals for sensitivity and specificity and no difference in median Ct values for any assay target between laboratories. Ct values were significantly lower in patients ≥ 45 years (*p* = 0.008).

**Conclusions and Significance:** PANDAA^®^ LASV demonstrated high diagnostic accuracy to detect Lassa virus RNA compared to the dual-reaction reference assay. Reducing the operational burden from two PCR reactions per sample to a single PANDAA^®^ LASV reaction while maintaining equivalent sensitivity and specificity could have a direct impact on labour and reagent costs, and turnaround time. Clinical validation against additional LASV lineages is an immediate priority.

## INTRODUCTION

Lassa virus (*Mammarenavirus lassaense*; LASV) is the causative agent of Lassa fever (LF), which is endemic to West Africa, including Nigeria, Liberia, Guinea, and Sierra Leone, where 100-300,000 infections, and 5,000 LASV-associated deaths, occur each year.^1^ National surveillance by the Nigeria Centre for Disease Control and Prevention (NCDC) reported 833–1,148 confirmed cases annually between 2019 and 2025, with case fatality rates of 16.3–27%.^2–7^ However, the true incidence is likely much higher as serology studies in endemic areas indicate that most infections go unreported with only ∼20% of Lassa fever cases reported with many infections asymptomatic or not captured by surveillance systems.^8–11^ Primarily transmitted by *Mastomys natalensis* rodents,^12^ human-to-human Lassa virus transmission in community or hospital settings remains a serious public health concern.^13^

The only widely used treatment for LASV infection is the broad-spectrum antiviral ribavirin, which decreases mortality rates when administered within the first six days of symptom onset.^14,15^ As LF typically presents with non-specific symptoms that overlap with other endemic febrile illnesses, insufficient diagnostic infrastructure forces clinicians to rely on differential diagnosis that can lead to initial treatment with antimalarials or antibiotics.^16^ LASV infection is suspected if symptoms do not resolve, and this delay reduces the likelihood of effective ribavirin treatment and increases LASV transmission to patient visitors and healthcare workers.^16^ Severe LF can progress to the haemorrhagic phase with multi-organ failure and high mortality with case fatality rates ranging 15–20% in hospitalised patients.^17^ Pregnant women are at elevated risk and survivors may develop long-term sequelae such as sensorineural hearing loss.^18,19^

The World Health Organization (WHO) designated LF as a priority disease under the WHO Blueprint for Action to Prevent Epidemics and collaborative international efforts are ongoing to improve surveillance systems and determine more accurate estimations of LF disease prevalence and LASV infection incidence.^20^ Although RT-PCR assays are the current standard for LF diagnosis due to their high analytical sensitivity and specificity, the high genetic diversity of LASV poses significant challenges.^21^ LASV groups into seven lineages that cluster geographically: lineages I–III in Nigeria, lineage IV in West Africa (Guinea, Sierra Leone, and Liberia),^22^ and lineage V in Mali and Côte d’Ivoire.^23^ Lineages VI and VII were identified more recently in Togo and Benin. With inter-lineage nucleotide variability ranging 19-27%,^24^ sequence variation at primer- and probe-binding sites, which can reduce PCR efficiency and impair diagnostic sensitivity, has made it difficult to develop a universal molecular diagnostic that captures all viral variants using conventional realtime PCR design.

The current diagnostic standard in Nigeria is the RealStar^®^ Lassa Virus RT-PCR Kit 2.0 (altona Diagnostics Germany) comprising two RT-PCR reactions targeting separate genomic regions (GPC and L gene) that are run simultaneously on each patient sample.^25^ This dual-reaction approach is designed to mitigate false negatives associated with viral variability. Yet it also highlights the inability of any single target conventional RT-PCR assay to reliably detect all LASV lineages and exposes significant weaknesses in the current diagnostic protocols as testing expands in West Africa, emphasising the need for pan-LASV PCR tests to address genetic variability and improve diagnostic reliability.^26^

The PANDAA^®^ LASV assay was developed to address the limitations of conventional real-time PCR using Pan-Degenerate Amplification and Adaptation (PANDAA) technology to detect all LASV lineages with high sensitivity and specificity.^27^ Unlike conventional real-time PCR, PANDAA overcomes known intra- and inter-lineage genetic variation while adapting to nucleotide diversity in novel viral variants in a single reaction. PANDAA has been used previously for HIV drug resistance genotyping.^28–30^ This is the first clinical evaluation of PANDAA LASV in an endemic setting. We conducted a diagnostic accuracy study comparing PANDAA^®^ LASV with the RealStar® Lassa Virus RT-PCR Kit 2.0 in Nigerian patients presenting with suspected Lassa fever at two sites.

## METHODS

### Ethics statement

The study was approved by the Research & Ethics Committee ATBUTH, Bauchi (ATBUTH/ADM/42/VOL.I), and the FMCO Health Research Ethics Committee (FMC/OW.380/VOL.LXXXII/21). Approval was given by both committees for the use of de-identified demographic, clinical, and laboratory diagnostic data that had been collected as part of routine clinical practice. Individual consent was not required as samples were collected for routine diagnostic purposes and excess clinical samples used for this study were obtained under ethics approval and de-identified prior to testing with PANDAA LASV.

### Study location and participants

The study was conducted at two diagnostic laboratories in Nigeria: Abubakar Tafawa Balewa University Teaching Hospital in Bauchi (ATBUTH Bauchi) (predominantly LASV lineage III), and Federal Medical Centre (FMC) Owo in Ondo (LASV lineage II).^31^ We used deidentified human EDTA plasma samples collected from patients evaluated for suspected Lassa fever. Available patient data from the testing request form included age, gender, and the number of days between symptom onset and sample collection. Patients were identified by the treating clinician based on the Nigeria Centre for Disease Control case definition for suspected Lassa fever, which includes acute febrile illness (≥ 38 °C) unresponsive to antimalarials or antibiotics, with or without hemorrhagic manifestations, in a patient with epidemiological risk factors. No adverse events were associated with this study as all testing was performed on excess clinical specimens collected during routine care. Alternative diagnoses were not systematically recorded for patients who tested negative for Lassa virus RNA as samples were collected under routine fever surveillance and diagnostic follow-up data were not available for this study.

### Routine laboratory testing and reference assay

As part of routine clinical care, EDTA plasma samples had been collected from patients meeting the standard case definition of suspected Lassa fever for testing by real-time PCR.^25,32^ Lassa virus RNA detection for routine diagnosis was performed using the RealStar^®^ Lassa Virus RT-PCR kit 2.0 (abbreviated here as the Altona assay), which was selected as the comparator assay because it is the reference standard deployed across Lassa fever diagnostic laboratories in Nigeria and the basis on which clinical management decisions are made at both study sites.^25^ The Altona assay was performed according to the manufacturer’s instructions and is two separate real-time PCR reactions for GPC and L viral genes run for 45 cycles. Lassa fever diagnostic laboratories in Nigeria use a positivity cut-off of Ct ≤ 42 cycles for the Altona assay, as no universal Ct threshold exists for Lassa virus RT-PCR.^25^ Amplification curves were visually inspected as part of routine practice to confirm that reported positives reflected true exponential amplification.^32^

### Study design

We compared PANDAA LASV with the Altona assay in a two-part diagnostic accuracy study (**Figure 1**). The overall enrolment target was 300 samples split equally between sites, with 150 positive and 150 negative specimens anticipated from the combined retrospective and prospective arms. The first was a retrospective diagnostic cohort study that tested 100 samples per site (*n* = 200) from specimens collected between January and April 2023. Sites were instructed to select approximately 70 positive and 30 negative samples, based on their positivity for Lassa virus RNA as determined by the Altona assay, to enrich the retrospective cohort for confirmed LASV-positive samples (enriched convenience sampling). As sample selection for the retrospective arm was based on the known Altona result, operators were not blinded to the reference standard classification when testing with PANDAA LASV. A target of 140 positive retrospective samples across both sites was selected to provide ≥ 95% power: if the assay detected 133/140 positives (point estimate 95.0%), the exact 95% CI would be 90.0–98.0%. Sixty negative retrospective samples per site would yield a 95% CI of 91.1– 100.0% for ≥ 98% specificity (59/60 negative). The prospective arm was expected to yield a positivity rate of approximately 10% based on historical data from the Nigeria Centre for Disease Control. Combined across both arms, the enrolment target was 150 positive and 150 negative samples. At this sample size, if the assay detected 143/150 positives (point estimate 95.3%), the exact 95% CI would be 90.6–98.1%; for specificity, 150 negative samples would yield a 95% CI of 94.3–99.6% at ≥ 98% specificity (147/150).

**Figure 1.**
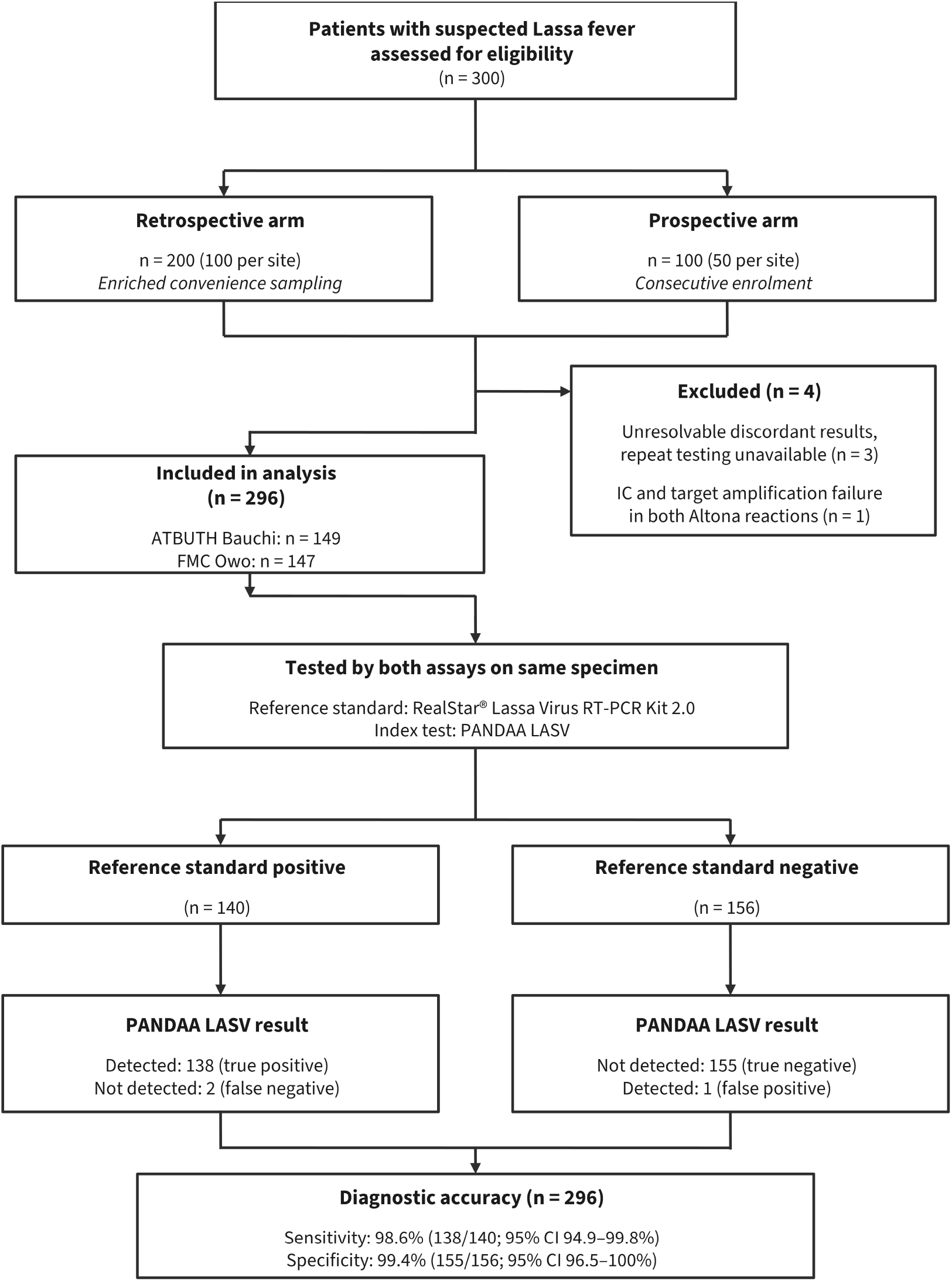
STARD flow diagram of participant enrolment and sample disposition.

To be eligible for the retrospective arm, plasma samples were required to have ≥ 500 µL available volume to allow for repeat testing and had undergone no freeze-thaw cycles since storage at −80 °C. RNA was freshly extracted from stored plasma and tested with PANDAA LASV between June and July 2023. The Altona assay had already been performed on these samples as part of routine clinical care and was only repeated if results were discordant with PANDAA. Both assays were performed on RNA extracted from the same plasma specimen for each patient; no clinical interval separated the two tests. For retrospective samples, PANDAA LASV was performed on freshly extracted RNA from plasma that had remained at −80 °C since collection.

A prospective diagnostic cohort study included 50 samples per site (*n* = 100) between June and July 2023. Each prospective sample was extracted and the eluted RNA tested first with the Altona assay, then with PANDAA LASV, in the order in which specimens were presented to the laboratory (consecutive enrolment). All prospective samples were tested by both assays regardless of the Altona result; there was no selection or enrichment based on Altona status. As the Altona assay had been performed on all samples prior to enrolment in the study (retrospective arm) or prior to PANDAA testing (prospective arm), the reference standard classification was made independently of the PANDAA result at both sites.

Viral RNA was from 100 µL plasma extracted using QIAamp Viral RNA Mini Kit (Qiagen, Germany) according to manufacturer’s instructions with nucleic acid eluted in 60 µL. Extracted RNA was used immediately for real-time PCR testing. Discordant results were resolved by re-extracting RNA from the stored plasma and repeating both Altona and PANDAA LASV assays.

### PANDAA LASV real-time PCR

PANDAA LASV was performed according to the manufacturer’s instructions with 10 µL eluted RNA used in a single 20 µL total reaction volume. PANDAA LASV was performed on a Mic Real Time PCR Cycler (Bio Molecular Systems) at ATBUTH Bauchi and a Rotor-Gene Q (Qiagen) at FMC Owo. The Altona assay was performed on the same instrument as PANDAA LASV at each site. PANDAA LASV cycling conditions were: 50 °C for 15 minutes, 95 °C for 2 minutes, followed by 10 adaptation cycles of 90 °C for 3 seconds, 55 °C for 30 seconds and 60 °C for 30 seconds. Amplification and detection occurred over 30 cycles of 90 °C for 3 seconds and 60 °C for 60 seconds. Fluorescence for LASV was acquired in the green channel and the internal control in the yellow channel.

PANDAA LASV contains an exogenous, non-competitive extraction control (Internal Control) that is spiked into the lysis buffer prior to extraction. High Lassa virus viral load may bias consumption of reaction amplification components and cause a delayed or absent Internal Control signal. Detection of the Internal Control is therefore not required to call a positive LASV result. For LASV-negative samples, the Internal Control must be detected; failure to detect is reported as an invalid result.

Each run included a Positive Control (acceptance criterion: Ct ≤ 25) and a Negative Control (acceptance criterion: undetected). At ATBUTH Bauchi, retrospective samples were tested across four runs and prospective samples across two runs on the Mic. At FMC Owo, retrospective samples were tested across three runs and prospective samples across two runs on the Rotor-Gene Q. Runs for discordant repeat testing are excluded from these totals.

For the Rotor-Gene Q, analysis settings were a threshold of 0.01 using dynamic tube normalisation for background removal with slope correct enabled. For the Mic, analysis settings were a threshold of 1.0 using the dynamic normalisation for background removal method with simple exclusion enabled to exclude samples with insufficient exponential amplification from Ct calling. As the 10 adaptation cycles do not capture fluorescence data, the PANDAA LASV positivity cut-off is Ct ≤ 30 to reflect data captured during the 30 amplification and detection cycles.

### Site training and qualification

Prior to study initiation, Aldatu Biosciences trained a laboratory operator from Market Access Africa, an independent consulting firm contracted to facilitate the study. This operator subsequently trained laboratory staff at each site. Aldatu provided each site with PANDAA LASV Training Kits containing ten samples prepared from non-infectious synthetic RNA covering the target regions of the Lassa virus genome. Seven samples represent the seven known LASV lineages at 50 cp/rxn (5× the assay limit of detection). Two additional samples at 5.0 × 10^4^ and 5.0 × 10^3^ copies/reaction form a standard curve with the 50 cp/rxn sample and the Positive Control to verify amplification efficiency and instrument performance. LCMV at 5.0 × 10^3^ copies/reaction is included as a specificity control, as LCMV is a mammarenavirus closely related to LASV. Laboratory operators ran the Training Kit before the study began, and results were returned to Aldatu to confirm accuracy, replicate consistency, and instrument qualification.

### Data analysis

A sample was called as positive for Lassa virus RNA using the Altona assay if either the GPC or L gene target was detected with a Ct ≤ 42. For PANDAA LASV, which is a single-reaction assay, the positivity cut-off was Ct ≤ 30. Samples returning no amplification of both the PANDAA LASV target and internal control were classified as invalid and excluded from analysis. Discordant results between the two assays were resolved by re-extraction and repeat testing of both assays where sufficient sample volume remained; samples that could not be resolved were excluded. Samples with missing demographic data were included in the diagnostic accuracy analysis but excluded from the relevant subgroup comparison; fluctuating denominators in demographic tables reflect this approach.

Sensitivity, specificity, accuracy, positive predictive value, and negative predictive value were calculated from 2 × 2 contingency tables with the Altona result as the reference classification; 95% confidence intervals for these measures are exact Clopper-Pearson intervals. Positive and negative diagnostic likelihood ratios and their 95% confidence intervals were calculated using the log method.^33^ Correlations between PANDAA and Altona Ct values were assessed using Spearman’s rank correlation coefficient. The Mann-Whitney rank-sum test was used to compare medians and other continuous variables between groups, and the Fisher’s exact test was used for categorical variables. Associations were considered statistically significant at *p* < 0.05. Diagnostic accuracy (sensitivity, specificity, PPV, NPV, likelihood ratios) and between-site comparisons were pre-specified analyses. Associations between patient demographics (age, gender) and Ct values were exploratory. All statistical analyses were performed in GraphPad Prism v8.4.3 and data were reported in accordance with the STARD-guidelines.^34^

## RESULTS

Three hundred samples from patients with suspected Lassa fever were tested across the two study sites with each site contributing 100 retrospective and 50 prospective samples. For the retrospective arm, sites were instructed to select approximately 70 positive and 30 negative samples (*n* = 200) to ensure sufficient power for sensitivity estimation, as the prospective arm was expected to yield a low positivity rate, which was determined to be 16% (**Table 1**). The final retrospective sample selections were 60 positive and 40 negative samples at ATBUTH Bauchi and 67 positive and 33 negative samples at FMC Owo (**Table 1**) from specimens collected between January and April 2023 that were previously tested using the RealStar® Lassa Virus RT-PCR kit. The prospective arm enrolled 50 consecutive samples per site (*n* = 100) between June and July 2023 and were tested by both assays without prior knowledge of the results from the Altona assay. Four samples were excluded from further analysis: at FMC Owo, three retrospective samples could not be resolved as repeat testing was unavailable for discordant results and at ATBUTH Bauchi, one prospective sample failed amplification of both the internal control and the Lassa target in both Altona reactions. The final cohort comprised 296 samples (ATBUTH Bauchi, *n* = 149; FMC Owo, *n* = 147).

**Table 1.** Distribution of samples by study site and study design. Data show the sample distribution after exclusion for unresolvable discordant samples. Four samples were excluded: three retrospective samples at ATBUTH Bauchi with discordant results that could not be resolved as repeat testing was unavailable and one prospective sample that failed amplification of the internal control and the Lassa targets in both Altona assay reactions.

| Site | Retrospective |  | Prospective |  | Total |  |
|---|---|---|---|---|---|---|
|  | Positive | Negative | Positive | Negative | Positive | Negative |
| ATBUTH Bauchi | 60 | 40 | 9 | 40 | 69 | 80 |
| FMC Owo | 64 | 33 | 7 | 43 | 71 | 76 |
| Combined (n = 296) | 124 | 73 | 16 | 83 | 140 | 156 |

### Diagnostic performance

From the remaining samples, 140/296 samples were positive for Lassa virus RNA by the Altona assay after retesting to resolve for discordant results (**Table 1**). PANDAA LASV detected 138/140 positive samples (**Supplementary Table 1**). Of the two positive samples that were negative by PANDAA, one was positive by L gene only (Ct = 37.7) and the other was positive by both Altona targets at high Ct values (GPC Ct= 41.1; L Ct= 38.9), close to the Altona cut-off of 42 cycles (**Supplementary Table 3**). Of the 156 Altona-negative samples, PANDAA LASV returned one discordant positive result. This sample had detectable amplification by both Altona targets (GPC Ct = 42.5; L Ct= 42.8) but was called negative as both values exceeded the 42-cycle cut-off.

PANDAA LASV sensitivity was 98.6% (138/140; 95% CI 94.9–99.8%), specificity 99.4% (155/156; 95% CI 96.5–100%), and accuracy 99.0% (293/296; 95% CI 97.1–99.8%) (**Table 2**). The positive diagnostic likelihood ratio exceeded 20 at the lower confidence bound (153.8; 95% CI 21.8–1084.9), and the negative likelihood ratio was 0.01 (95% CI 0.00–0.06). Performance was consistent at both sites: sensitivity was 97.1% (67/69) at ATBUTH Bauchi and 100% (71/71) at FMC Owo, and specificity was 100% (80/80) and 98.7% (75/76), respectively.

**Table 2.** PANDAA LASV performance. Site-specific likelihood ratios were not calculated; zero false positives at ATBUTH Bauchi and zero false negatives at FMC Owo preclude stable estimation.

|  | **ATBUTH Bauchi** | ***n*** | **FMC Owo** | ***n*** | **Combined (n=296)** | ***n*** |
|---|---|---|---|---|---|---|
| Sensitivity | 97.1% (89.9–99.6%) | 67/69 | 100% (94.9–100%) | 71/71 | 98.6% (94.9–99.8%) | 138/140 |
| Specificity | 100% (95.5–100%) | 80/80 | 98.7% (92.9–100%) | 75/76 | 99.4% (96.5–100%) | 155/156 |
| Accuracy | 98.7% (95.3–99.8%) | 147/149 | 99.3% (96.3–100%) | 146/147 | 99.0% (97.1–99.8%) | 293/296 |
| PPV | 100% (94.6–100%) | 67/67 | 98.6% (92.5–100%) | 71/72 | 99.3% (96.1–100%) | 138/139 |
| NPV | 97.6% (91.5–99.7%) | 80/82 | 100% (95.2–100%) | 75/75 | 98.7% (95.5–99.8%) | 155/157 |
| Positive LR | — | — | — | — | 153.8 (21.8–1084.9) | — |
| Negative LR | — | — | — | — | 0.01 (0.00–0.06) | — |

### Discordant results and Ct value distributions

Prior to repeat testing, 18/296 samples (6.1%) were discordant (**Supplementary Table 2**). Sixteen samples that were originally positive by Altona and negative by PANDAA, 14 (87.5%) were found to be negative by Altona upon repeat testing (**Supplementary Table 2**). Of those, 9/14 (64.3%) had only one of the two Altona target genes detected in the original real-time PCR run with a median Ct of 38.0 cycles (IQR: 35.4–39.4; **Supplementary Table 4**). Two samples were positive by PANDAA and negative by Altona and repeat testing returned one concordant positive result by the Altona assay (L gene Ct= 38.8).

The median Ct value for samples positive by PANDAA was 23.2 (IQR: 18.7–26.1). Median Ct values for the Altona GPC and L gene targets were 31.4 (IQR 27.3–35.9) and 32.0 (IQR 26.0–34.8), respectively. Note that PANDAA acquires fluorescence over 30 cycles following a 10-cycle adaptation phase that does not generate signal; the Altona assay acquires over 45 cycles. There was no difference in median Ct observed between study sites for any assay target (**Table 3**). Ct values for the PANDAA true positive samples (*n* = 138) were correlated with the Altona assay where the GPC and/or L gene targets returned a Ct value (**Figure 2**). Significant, positive correlations were observed between PANDAA Ct values and Altona GPC Ct values (Spearman’s *ρ* = 0.69; 95% CI 0.58–0.77; *p* < 0.0001; *n* = 135) and between PANDAA and Altona L gene Ct values (*ρ* = 0.80; 95% CI 0.73–0.86; *p* < 0.0001; *n* = 133). Altona GPC and L Ct values were also positively correlated (*ρ* = 0.84, 95% CI 0.78–0.88; *p* < 0.0001) among samples with both Altona targets detected (*n* = 131).

**Table 3.** Ct value distributions for Lassa-positive samples by study site.

|  | ATBUTH Bauchi |  | FMC Owo |  |  | Combined |  |
|---|---|---|---|---|---|---|---|
| Assay (target) | Median Ct (IQR) | Ct Range | Median Ct (IQR) | Ct Range | *p* value | Median Ct (IQR) | Ct Range |
| PANDAA | 22.8 (17.8–25.3) | 8.1–29.5 | 23.7 (19.6–26.6) | 7.0–30.0 | 0.113 | 23.2 (18.7–26.1) | 7.0–30.0 |
| Altona (GPC) | 31.4 (26.2–36.5) | 17.8–41.1 | 31.5 (29.2–35.2) | 20.7–39.9 | 0.977 | 31.4 (27.3–35.9) | 17.8–41.1 |
| Altona (L) | 31.7 (24.7–34.8) | 18.0–40.5 | 32.6 (28.0–34.8) | 22.6–39.0 | 0.220 | 32.0 (26.0–34.8) | 18.0–40.5 |

**Figure 2.**
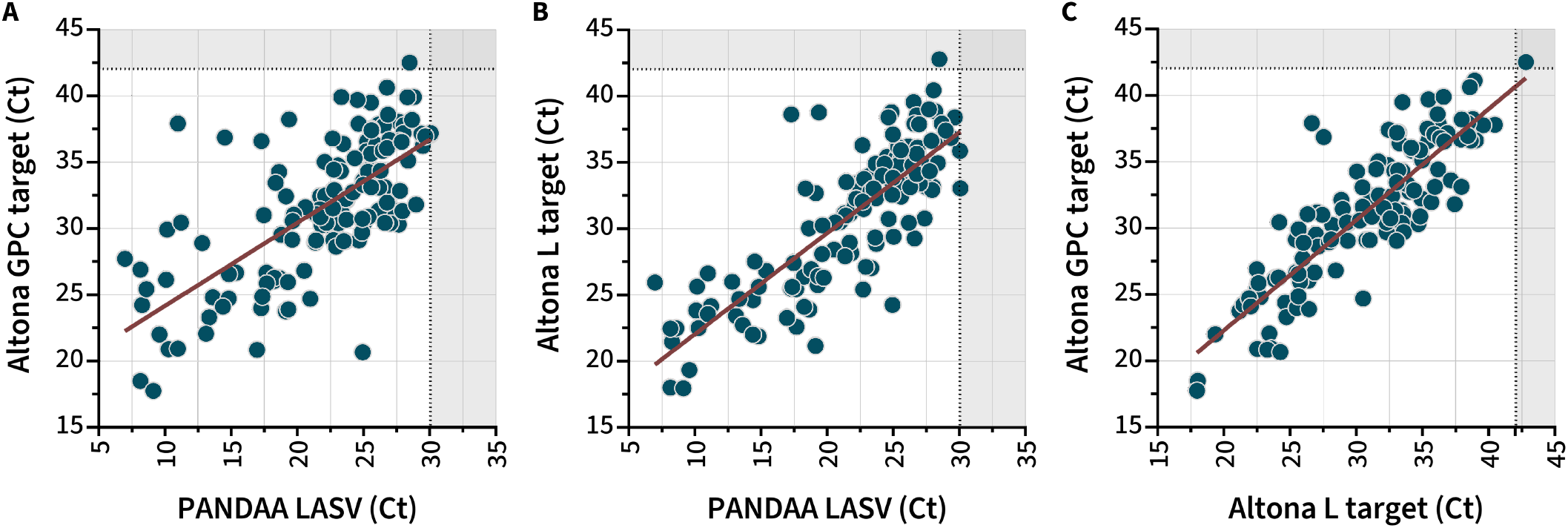
Ct correlations between the PANDAA LASV and Altona assay targets. Spearman’s correlation coefficients (*ρ*) were calculated using only the Ct values that were detectable for both assays in the pair. Grey shaded areas indicate the assays cut-off Ct values of 30 cycles for PANDAA and 42 for the Altona targets. (**A**) correlation between paired PANDAA and Altona GPC target Ct values. (*ρ* = 0.69; 95% CI: 0.58–0.77; *p* < 0.0001) (*n* = 135). Three of 138 PANDAA true positives lacked a GPC Ct value and were excluded. One sample is shown above the Altona GPC assay cut-off, which is the discordant positive result by PANDAA. (**B**) correlation between PANDAA and the Altona L gene target (*ρ* = 0.80; 95% CI: 0.73–0.86; *p* < 0.0001) (*n* = 133). Five of 138 PANDAA true positives lacked an L gene target Ct value and were excluded. The discordant positive result by PANDAA is shown above the Altona L gene cut-off (**C**) correlation between the two Altona assay targets. (*ρ* = 0.84; 95% CI: 0.78–0.88; *p* < 0.0001) (*n* = 131). Nine of 140 Altona-positive samples had only one gene target detected and were excluded. The discordant positive result by PANDAA is identifiable by the single sample shown above the cut-off for both Altona assays.

### Patient characteristics

The characteristics of the 140 Lassa fever positive individuals evaluated in this study are presented in **Table 4**. There was no significant difference in gender distribution between the two sites for samples tested by PANDAA LASV retrospectively, and in total 44.6% were female. Overall, patients ranged in age from 0 to 88 years with a median of 34 (IQR: 21.0–48.0) of which 15.0% were children under the age of 18 years (*n* = 21) with no difference in the number of children between the two sites (*p* = 0.347, Fisher’s exact test). However, there was a significant difference in patient age in retrospective samples, with a median age of 28 years at ATBUTH Bauchi compared to 37 years at FMC Owo (*p* = 0.0074, Mann-Whitney). There was also a significant difference in the time between symptom onset and sample collection, with a median of 3.5 days (IQR 1–7) at ATBUTH Bauchi and 7 days (IQR 4–9) for samples tested at FMC Owo (*p* < 0.001, Mann-Whitney). Although we found no association between gender and Ct value (**Supplementary Table 5**), those over 45 years of age had significantly lower Ct values using the PANDAA LASV assay (*p* = 0.008, Mann-Whitney test) and for the GPC target of the Altona assay (*p* = 0.003, Mann-Whitney test)(**Supplementary Table 6**).

**Table 4.** Characteristics of Lassa fever positive cases. Sociodemographic characteristics of Lassa PCR-positive patients included in the comparison (*n* = 140). Data are given as *n* (%), or median (IQR). Missing data is indicated in the table by the fluctuating denominator (*n*).

|  | Retrospective Positive Samples (*n*=124) |  | Prospective Positive Samples (*n*=16) |  |  |
|---|---|---|---|---|---|
|  | ATBUTH Bauchi | FMC Owo | ATBUTH Bauchi | FMC Owo | Total |
| **Gender**, *n* (%) |  |  |  |  |  |
| Female | 22 (37.3%) | 34 (53.1%) | 2 (22.2%) | 4 (57.1%) | 62 (44.6%) |
| Male | 37 (62.7%) | 30 (46.9%) | 7 (77.8%) | 3 (42.9%) | 77 (55.4%) |
| **Age**, median years (IQR) | 29 (20.0–40.0) | 39.5 (24.0–53.8) | 30.0 (20.0–35.0) | 45.0 (44.0–57.5) | 34 (21.0–48.0) |
| **Age group**, years |  |  |  |  |  |
| < 10 | 3 (5.0%) | 5 (7.8%) | 0 (0.0%) | 1 (11.1%) | 9 (6.4%) |
| 10–19 | 6 (10.0%) | 8 (12.5%) | 2 (22.2%) | 0 (0.0%) | 16 (11.4%) |
| 20–29 | 21 (35.0%) | 11 (17.2%) | 2 (22.2%) | 0 (0.0%) | 34 (24.3%) |
| 30–39 | 14 (23.3%) | 8 (12.5%) | 3 (33.3%) | 0 (0.0%) | 25 (17.9%) |
| 40–49 | 6 (10.0%) | 12 (18.8%) | 1 (11.1%) | 3 (33.3%) | 22 (15.7%) |
| 50–59 | 6 (10.0%) | 6 (9.4%) | 0 (0.0%) | 1 (11.1%) | 13 (9.3%) |
| 60–69 | 3 (5.0%) | 9 (14.1%) | 1 (11.1%) | 1 (11.1%) | 14 (10.0%) |
| > 69 | 1 (1.7%) | 5 (7.8%) | 0 (0.0%) | 1 (11.1%) | 7 (5.0%) |
| **Onset to sampling**,<br>median (IQR) (*n*) | 2.5 (1–6) days<br>(44/60) | 6 (4–8) days<br>(64/64) | 1 (1–1) days<br>(9/9) | 7 (6–8) days<br>(7/7) | 5 (1–7) days<br>(124/140) |

## DISCUSSION

In this head-to-head comparison against the RealStar Lassa Virus RT-PCR kit 2.0 at two Nigerian Lassa fever diagnostic laboratories, PANDAA LASV detected Lassa virus RNA in a single RT–PCR reaction with sensitivity of 98.6% and specificity of 99.4%. Site-level performance was similar: sensitivity was 97.1% at ATBUTH Bauchi (lineage III) and 100% at FMC Owo (lineage II); specificity was 100% and 98.7%, respectively. The positive likelihood ratio of 153.8 (95% CI 21.8–1084.9) and negative likelihood ratio of 0.01 (95% CI 0.00–0.06) indicate that a PANDAA result carries high diagnostic weight in either direction. To our knowledge, this is the first clinical evaluation of a pan-lineage, single-reaction Lassa virus RT–PCR assay in an endemic setting.

Of the three confirmed discordant results, two were negative by PANDAA in samples with high Ct values by the Altona assay: one positive by L gene alone (Ct = 37.7) and the other by both targets near the 42-cycle cutoff (GPC 41.1, L 38.9). The third discordant result was positive by PANDAA and negative by the Altona assay, with both Altona assay targets 0.5–0.8 cycles above the cut-off. A limitation of this study was that a third assay was not available to independently verify the accuracy of these discordant results, particularly the single PANDAA false positive result given the pan-lineage design of PANDAA, which mitigates sequence variation at primer- and probe-binding sites that would otherwise reduce the efficiency of conventional real-time PCR assays. Such high Ct values indicate stochastic low copy number sampling and the resultant run-to-run variability in RT–PCR assays at the molecular limits of detection and not a systematic failure of either assay. This interpretation is supported by the resolution of 14/16 (87.5%) initially discordant false negative results that were found to be LASV-negative when re-extracted and retested with the Altona assay. Nine of the 14 had only a single Altona target detected in the original run, with Ct values near the assay cutoff (median 38.0; IQR 35.4–39.4). It cannot be determined from these data alone whether this represents original false positives by the Altona assay or RNA degradation from storage and handling of retrospective specimens.

PANDAA LASV is a single 20-µL reaction for pan-lineage Lassa virus detection. The Altona assay requires two separate RT–PCR reactions per patient sample to safeguard against genetic variation at any one target causing a false negative result. This approach requires double the reagent consumption, thermocycler capacity, and hands-on time per specimen. During peak Lassa fever season, diagnostic laboratories face significant throughput constraints. Reducing the operational burden from two PCR reactions per sample to a single PANDAA LASV reaction while maintaining equivalent diagnostic accuracy could have a direct impact on labour and reagent costs and turnaround time. The clinical evaluation in this study is limited to lineages II and III. The performance of PANDAA LASV against other lineages has only been confirmed using synthetic RNA constructs and has yet to be confirmed in the field with clinical samples.

Although Ct values differed between assays by approximately 8–9 cycles (PANDAA median 23.2 vs Altona GPC 31.4 and L 32.0), such a large offset is to be expected as a consequence of PANDAA assay design, which acquires fluorescence during the 30 amplification cycles and not during the preceding 10-cycle adaptation phase. Absolute Ct values between the two assays are not directly comparable, however, rank-order agreement was strong. PANDAA Ct correlated significantly with both Altona L gene Ct (*ρ* = 0.80; *p* < 0.0001) and Altona GPC (*ρ* = 0.69; *p* < 0.0001). The tighter correlation with the L gene target is to be expected as PANDAA LASV also targets the L segment, albeit at a distinct site.

The 140 Lassa-positive patients in this study were demographically comparable to previously reported Nigerian cohorts. Females accounted for 44.6%, similar to the LASCOPE prospective cohort across four Nigerian states between 2018 and 2019.^25^ The median age of 34 years was also similar to several documented studies,^25,32,35,36^ as was the proportion of children under 18 years (15.0%).^25^ It was notable that the median age was lower at ATBUTH Bauchi than FMC Owo (28 vs 37 years; *p* = 0.0074) although the design of this study cannot determine whether site-specific differences in age distribution reflect differences in the local population, referral patterns, or epidemiological factors. The time from symptom onset to sample collection was also shorter at ATBUTH Bauchi (median 3.5 vs 7 days; *p* < 0.001). However, this shorter onset-to-sampling interval at ATBUTH Bauchi did not produce a detectable difference in median Ct values between sites for any assay target. Patients over 45 years had significantly lower PANDAA Ct values than younger patients (*p* = 0.008) and Altona GPC target Ct values (*p* = 0.003) indicating higher viral loads. This observation is exploratory as this study was not powered for an age-stratified viral load analysis. This trend of a higher proportion of low Ct values in patients ≥ 45 years has been reported previously along with a higher case fatality rate in that age group with both variables independently predicting mortality.^15,25^

Pan-lineage detection has been a central challenge for Lassa virus molecular diagnostics. PANDAA LASV analytical validation demonstrated an assay-level limit of detection (LoD) of 10 RNA copies/reaction with all seven recognised LASV lineages and a systemlevel LoD for the end-to-end workflow (extraction → amplification) of 203 IU/mL (95% CI: 146–283 IU/mL) using the First WHO International Standard for Lassa virus (LASV) RNA (NIBSC 21/112; Lineage IV, Josiah).^37^ In a 2025 WHO collaborative study across 18 international laboratories using chimeric lentiviral particles for five LASV lineages, PANDAA LASV was the only commercially available single reaction real-time PCR assay to detect all blinded samples.^37^ Approximately half of the single-target assays in that evaluation failed to detect lineage III (8/14) and lineage VII (7/14).

These gaps demonstrate why a diagnostic that can accommodate sequence variation by design, rather than by adding targets, has practical value. However, that value is only fully realised when confirmed against clinical specimens for each lineage. Implementing centralised molecular diagnostics that can reliably detect all circulating lineages is a critical prerequisite for the validation of rapid diagnostic tests (RDTs) for point-of-care Lassa fever detection. The field evaluation by Elsinga et al. of the ReLASV Pan-Lassa Antigen RDT demonstrated the critical nature of selecting the correct reference standard in RDT evaluations as an RDT’s measured sensitivity will reflect any insufficiencies in the reference assay used to define LASV-positive status.^35,38^ Accurate determination of LASV infection in vaccine trial participants, such as those by IAVI and the CEPI-funded Lassa virus vaccine efficacy trials in phase 2 enrolment in Nigeria, Ghana, Liberia, and Sierra Leone during 2024,^39,40^ also requires an assay with confirmed pan-lineage sensitivity and high specificity against related arenaviruses, such as LCMV.

This study had several limitations. No independent third assay was available to resolve discordant results, and the clinical evaluation is limited to LASV lineages II and III. The retrospective arm used enriched convenience sampling, and operators were not blinded to the Altona result when testing with PANDAA LASV. However, Ct values were generated by instrument software using predefined analysis thresholds, which limited observer-dependent bias in result interpretation, and the prospective arm enrolled consecutive samples tested by both assays from the same extraction eluate. Sample sizes were close to expected for positive and negative specimens, resulting in narrow 95% confidence intervals; however, the prospective arm was not independently powered for sensitivity estimation, with only 16 positive samples. The study was not designed to evaluate clinical outcomes or the prognostic value of PANDAA Ct values.

Confirming PANDAA LASV clinical performance in endemic regions with other circulating LASV lineages is an immediate priority. A multi-site study is needed to evaluate PANDAA LASV across a range of viral loads, lineages, and clinical presentations, while also capturing operational metrics (turnaround time, hands-on time per sample, and user feedback) to quantify the workflow advantages of a single-reaction format, in routine Lassa fever diagnostics.

## Data Availability

All data produced in the present work are contained in the manuscript

## SUPPLEMENTARY INFORMATION

**Supplementary Table 1.**
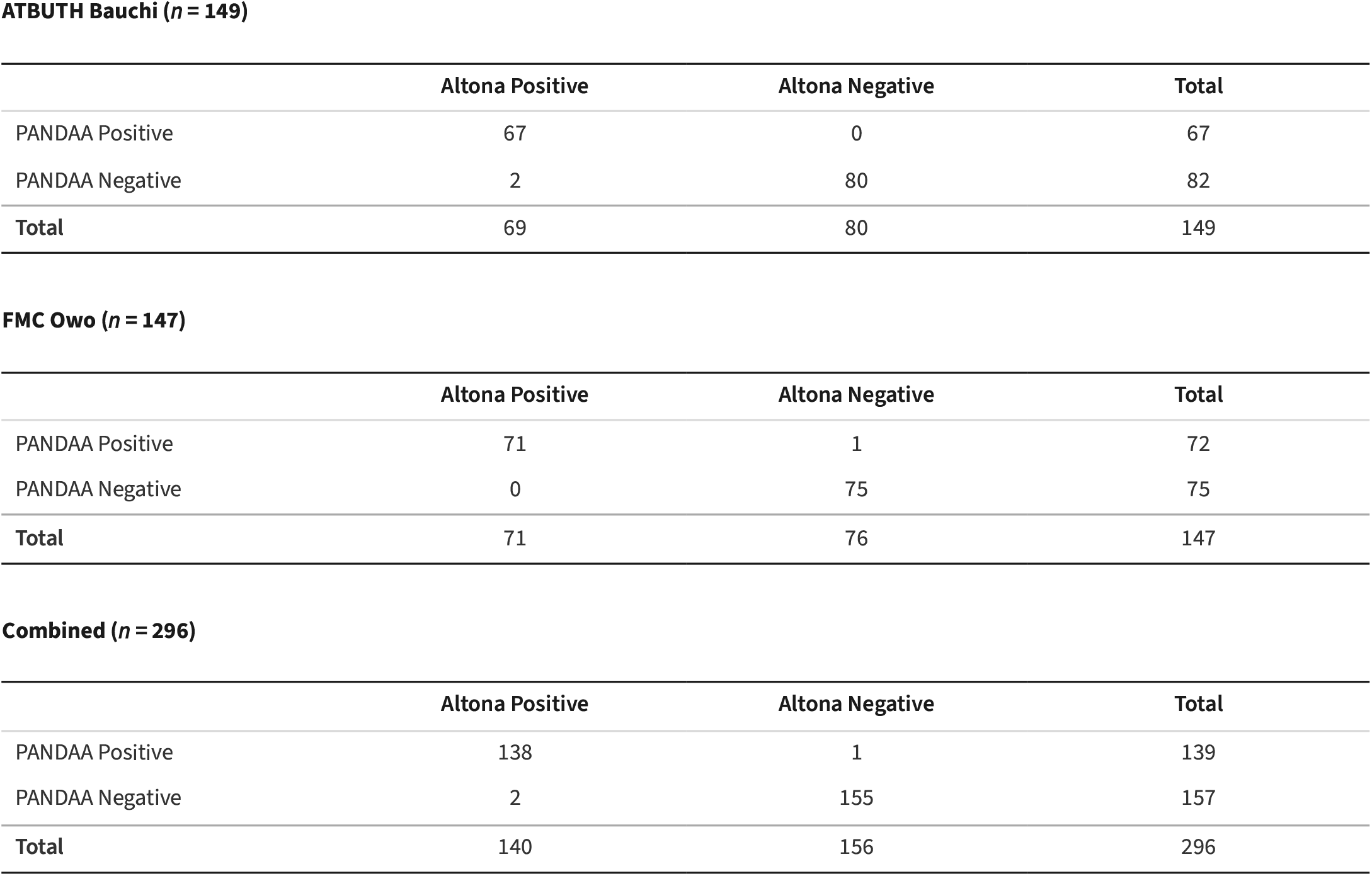
Cross-tabulation of PANDAA LASV results against the Altona reference classifications.

| **ATBUTH Bauchi (*n* = 149)** |  |  |  |
|  | **Altona Positive** | **Altona Negative** | **Total** |
|---|---|---|---|
| PANDAA Positive | 67 | 0 | 67 |
| PANDAA Negative | 2 | 80 | 82 |
| **Total** | 69 | 80 | 149 |
| **FMC Owo (*n* = 147)** |  |  |  |
|  | **Altona Positive** | **Altona Negative** | **Total** |
| PANDAA Positive | 71 | 1 | 72 |
| PANDAA Negative | 0 | 75 | 75 |
| **Total** | 71 | 76 | 147 |
| **Combined (*n* = 296)** |  |  |  |
|  | **Altona Positive** | **Altona Negative** | **Total** |
| PANDAA Positive | 138 | 1 | 139 |
| PANDAA Negative | 2 | 155 | 157 |
| **Total** | 140 | 156 | 296 |

**Supplementary Table 2.**
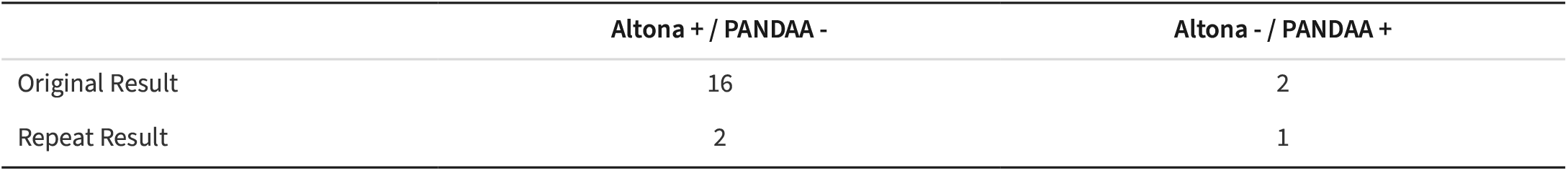
Resolution of discordant results.

|  | **Altona + / PANDAA -** | **Altona - / PANDAA +** |
|---|---|---|
| Original Result | 16 | 2 |
| Repeat Result | 2 | 1 |

**Supplementary Table 3.** Ct values of discordant results.

| Site | Sample ID | Altona Result | GPC Ct Value | L Ct Value | PANDAA Result | PANDAA Ct |
|---|---|---|---|---|---|---|
| Bauchi | ATBU/MOGID/LF/23/1751 | POSITIVE | 41.13 | 38.93 | NEGATIVE | N.D. |
| Bauchi | ATBU/MOGID/LF/23/2046 | POSITIVE | N.D. | 37.68 | NEGATIVE | N.D. |
| FMC Owo | FO/2018/LV/23 | NEGATIVE | 42.54 | 42.81 | POSITIVE | 27.68 |

**Supplementary Table 4.** Ct values of Altona results with single target (GPC or L) detected.

| Site | Sample ID | Altona Result | GPC Ct Value | L Ct Value | PANDAA Result | PANDAA Ct |
|---|---|---|---|---|---|---|
| Bauchi | ATBU/MOGID/LF/23/2020 | POSITIVE | N.D. | 33.15 | POSITIVE | 27.51 |
| Bauchi | ATBU/MOGID/LF/23/2046 | POSITIVE | N.D. | 37.68 | NEGATIVE | N.D. |
| Bauchi | ATBU/MOGID/LF/23/2153 | POSITIVE | N.D. | 38.88 | POSITIVE | 28.18 |
| Bauchi | ATBU/MOGID/LF/23/1789 | POSITIVE | N.D. | 38.81 | POSITIVE | 24.87 |
| Bauchi | ATBU/MOGID/LF/23/1917 | POSITIVE | 38.08 | N.D. | POSITIVE | 27.65 |
| Bauchi | ATBU/MOGID/LF/23/1926 | POSITIVE | 36.24 | N.D. | POSITIVE | 29.49 |
| Bauchi | ATBU/MOGID/LF/23/2181 | POSITIVE | 39.94 | N.D. | POSITIVE | 28.81 |
| Bauchi | ATBU/MOGID/LF/23/2217 | POSITIVE | 36.31 | N.D. | POSITIVE | 25.95 |
| Bauchi | ATBU/MOGID/LF/23/2265 | POSITIVE | 39.94 | N.D. | POSITIVE | 23.31 |

**Supplementary Table 5.** Association between gender and Ct value.

| **Assay (target)** | **Female**<br>**Median Ct (IQR)** | **Male**<br>**Median Ct (IQR)** | ***p* value \*** |
|---|---|---|---|
| PANDAA | 23.0 (18.6–25.6) | 23.7 (18.8–26.5) | 0.696 |
| Altona (GPC) | 31.4 (28.9–35.5) | 31.5 (27.5–36.8) | 0.526 |
| Altona (L) | 31.6 (26.3–34.9) | 32.8 (26.7–34.9) | 0.483 |

\* Mann-Whitney rank-sum test

**Supplementary Table 6.**
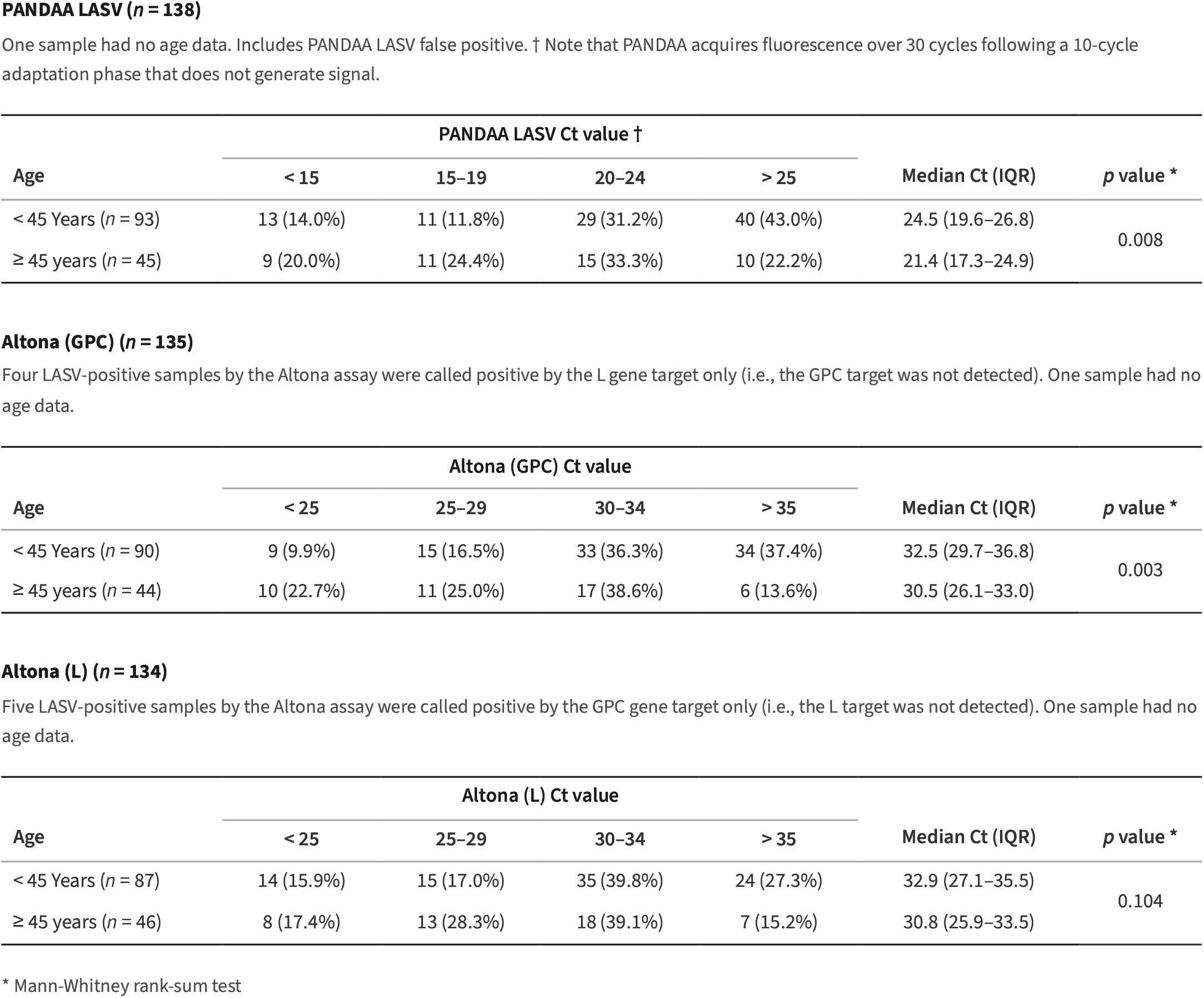
Distribution of Ct values by age.

| **PANDAA LASV (*n* = 138)** |  |  |  |  |  |  |
| One sample had no age data. Includes PANDAA LASV false positive. † Note that PANDAA acquires fluorescence over 30 cycles following a 10-cycle adaptation phase that does not generate signal. |  |  |  |  |  |  |
|  | **PANDAA LASV Ct value †** |  |  |  |  |  |
| **Age** | **< 15** | **15–19** | **20–24** | **> 25** | **Median Ct (IQR)** | ***p* value \*** |
|---|---|---|---|---|---|---|
| < 45 Years (*n* = 93) | 13 (14.0%) | 11 (11.8%) | 29 (31.2%) | 40 (43.0%) | 24.5 (19.6–26.8) | 0.008 |
| ≥ 45 years (*n* = 45) | 9 (20.0%) | 11 (24.4%) | 15 (33.3%) | 10 (22.2%) | 21.4 (17.3–24.9) |  |
| **Altona (GPC) (*n* = 135)** |  |  |  |  |  |  |
| Four LASV-positive samples by the Altona assay were called positive by the L gene target only (i.e., the GPC target was not detected). One sample had no age data. |  |  |  |  |  |  |
|  | **Altona (GPC) Ct value** |  |  |  |  |  |
| **Age** | **< 25** | **25–29** | **30–34** | **> 35** | **Median Ct (IQR)** | ***p* value \*** |
| < 45 Years (*n* = 90) | 9 (9.9%) | 15 (16.5%) | 33 (36.3%) | 34 (37.4%) | 32.5 (29.7–36.8) | 0.003 |
| ≥ 45 years (*n* = 44) | 10 (22.7%) | 11 (25.0%) | 17 (38.6%) | 6 (13.6%) | 30.5 (26.1–33.0) |  |
| **Altona (L) (*n* = 134)** |  |  |  |  |  |  |
| Five LASV-positive samples by the Altona assay were called positive by the GPC gene target only (i.e., the L target was not detected). One sample had no age data. |  |  |  |  |  |  |
|  | **Altona (L) Ct value** |  |  |  |  |  |
| **Age** | **< 25** | **25–29** | **30–34** | **> 35** | **Median Ct (IQR)** | ***p* value \*** |
| < 45 Years (*n* = 87) | 14 (15.9%) | 15 (17.0%) | 35 (39.8%) | 24 (27.3%) | 32.9 (27.1–35.5) | 0.104 |
| ≥ 45 years (*n* = 46) | 8 (17.4%) | 13 (28.3%) | 18 (39.1%) | 7 (15.2%) | 30.8 (25.9–33.5) |  |

\* Mann-Whitney rank-sum test

## FINANCIAL DISCLOSURE

Aldatu Biosciences, Inc. provided PANDAA^®^ LASV assay kits, PANDAA^®^ LASV Training Kits, and site training for this study and funded site-level study costs (labour, consumables, and logistics) through Market Access Africa, an independent consulting firm contracted to facilitate the study. No specific grant funding was received.

## COMPETING INTERESTS

Iain J. MacLeod is the co-founder, CEO, and Chief Scientific Officer of Aldatu Biosciences, Inc., which manufactures and commercialises PANDAA^®^ LASV. The PANDAA technology is covered by US Patent No. 10,100,349 and European Patent Application No. 3052656, owned by the President and Fellows of Harvard College and exclusively licensed to Aldatu Biosciences, Inc. Aldatu Biosciences funded the study as described in the Financial Disclosure. Market Access Africa, which was contracted and paid by Aldatu Biosciences to facilitate this study. All other authors declare no competing interests.

## AUTHOR CONTRIBUTIONS

**Conceptualization:** Iain J. MacLeod, Nancy Njoku-Attah

**Data curation:** Johnson Etafo, Idris I. Abduljalail, Ruth I. Aderibigbe, Aisha M. Yarima, Anita O. Atu, Olufunke Gbenga-Ayeni, Bawa A. Mikail, Ronke I. Uzuajemeh, Nelson A. Adedosu, Suleiman Y. Alhaji

**Formal analysis:** Iain J. MacLeod

**Funding acquisition:** Iain J. MacLeod, Nancy Njoku-Attah

**Investigation:** Johnson Etafo, Idris I. Abduljalail, Ruth I. Aderibigbe, Aisha M. Yarima, Anita O. Atu, Olufunke Gbenga-Ayeni, Bawa A. Mikail, Ronke I. Uzuajemeh, Nelson A. Adedosu, Suleiman Y. Alhaji

**Methodology:** Iain J. MacLeod, Nancy Njoku-Attah, Johnson Etafo, Nelson A. Adedosu, Suleiman Y. Alhaji

**Project administration:** Iain J. MacLeod, Nancy Njoku-Attah

**Resources:** Iain J. MacLeod, Nancy Njoku-Attah, Nelson A. Adedosu, Suleiman Y. Alhaji

**Supervision:** Nancy Njoku-Attah, Nelson A. Adedosu, Suleiman Y. Alhaji

**Writing – original draft:** Iain J. MacLeod

**Writing – review & editing:** Iain J. MacLeod, Nancy Njoku-Attah, Nelson A. Adedosu, Suleiman Y. Alhaji

